# Global disparities in SARS-CoV-2 genomic surveillance

**DOI:** 10.1101/2021.08.21.21262393

**Authors:** Anderson F. Brito, Elizaveta Semenova, Gytis Dudas, Gabriel W. Hassler, Chaney C. Kalinich, Moritz U.G. Kraemer, Joses Ho, Houriiyah Tegally, George Githinji, Charles N. Agoti, Lucy E. Matkin, Charles Whittaker, Danish Covid-19 Genome Consortium, COVID-19 Impact Project, Network for Genomic Surveillance in South Africa (NGS-SA), GISAID core curation team, Benjamin P Howden, Vitali Sintchenko, Neta S. Zuckerman, Orna Mor, Heather M Blankenship, Tulio de Oliveira, Raymond T. P. Lin, Marilda Mendonça Siqueira, Paola Cristina Resende, Ana Tereza R. Vasconcelos, Fernando R. Spilki, Renato Santana Aguiar, Ivailo Alexiev, Ivan N. Ivanov, Ivva Philipova, Christine V. F. Carrington, Nikita S. D. Sahadeo, Céline Gurry, Sebastian Maurer-Stroh, Dhamari Naidoo, Karin J von Eije, Mark D. Perkins, Maria van Kerkhove, Sarah C. Hill, Ester C. Sabino, Oliver G. Pybus, Christopher Dye, Samir Bhatt, Seth Flaxman, Marc A. Suchard, Nathan D. Grubaugh, Guy Baele, Nuno R. Faria

## Abstract

Genomic sequencing provides critical information to track the evolution and spread of SARS-CoV-2, optimize molecular tests, treatments and vaccines, and guide public health responses. To investigate the spatiotemporal heterogeneity in the global SARS-CoV-2 genomic surveillance, we estimated the impact of sequencing intensity and turnaround times (TAT) on variant detection in 167 countries. Most countries submit genomes >21 days after sample collection, and 77% of low and middle income countries sequenced <0.5% of their cases. We found that sequencing at least 0.5% of the cases, with a TAT <21 days, could be a benchmark for SARS-CoV-2 genomic surveillance efforts. Socioeconomic inequalities substantially impact our ability to quickly detect SARS-CoV-2 variants, and undermine the global pandemic preparedness.

**One-Sentence Summary:** Socioeconomic inequalities impacted the SARS-CoV-2 genomic surveillance, and undermined the global pandemic preparedness.

## The importance of genomic surveillance

Twenty months into the COVID-19 pandemic, many countries continue to face large epidemics of SARS-CoV-2 infections (*1*), mostly driven by the emergence and spread of novel viral variants (*2*). Genomic surveillance has been critical to the study of SARS-CoV-2 evolution and spread, to the design and optimization of diagnostic tools and vaccines, and to the early identification and assessment of viral lineages with altered epidemiological characteristics, including variants of concern (VOCs) such as Alpha/B.1.1.7; Beta/B.1.351; Gamma/P.1; and Delta/B.1.617.2. These lineages pose increased global public health risks due to their greater transmissibility and potential immune escape from neutralizing antibodies induced by natural infections and/or vaccines (*3, 4*). Variants of interest (VOIs) also require continued monitoring for changes in transmissibility, disease severity, or antigenicity (*5*). To help guide public health responses to evolving variants, it is essential to track the diversity of SARS-CoV-2 lineages circulating worldwide in near real-time (*3, 6, 7*). An unprecedented number of SARS-CoV-2 viral genomes have now been released in publicly accessible databases, with >4 million consensus genome sequences shared via the EpiCoV database at the GISAID data science initiative (*8*) and >1.2 million high-throughput sequencing datasets and >1.7 million consensus sequences in National Center for Biotechnology Information as of October 1^st^, 2021. Until then, and as a comparison, 324,992 influenza genome sequences have been shared in the GISAID database. Despite improvements in models for equitable sharing of pathogen genomic data (*9*), there are striking differences in the intensity of genomic surveillance within and among countries worldwide. Here we examine global publicly-accessible SARS-CoV-2 genomic surveillance data from the first 15 months of the COVID-19 pandemic to identify key aspects associated with sequencing intensity and timely variant detection, and investigate the consequences of surveillance disparities.

## Global disparities in SARS-CoV-2 genomic surveillance

To investigate spatial and temporal heterogeneity in SARS-CoV-2 genome sequencing intensity, we explored the percentage of COVID-19 cases sequenced each week per country from February 2020 to March 2021 (**Fig. 1A; Table S1**). It has been proposed that at least 5% of SARS-CoV-2 positive samples should be sequenced to detect viral lineages at a prevalence of 0.1 to 1.0% (*10*). Only 16 countries (or 9.6%) worldwide sequenced 5% or more of their total confirmed cases, while 100 out of 167 countries had <0.5% of confirmed cases sequenced (**Fig. 1B; Fig. S1**). A total of 72 countries had <25% of their genomes sequenced locally, and relied mostly on sequencing capacity in other countries to get their cases sequenced (**Fig. S2; Table S2**). Among high-income countries (HICs) and low- and middle-income countries (LMICs), while the number of reported cases was relatively similar until late March 2021 (65.3 and 61.2 million, respectively), HICs shared on average 16.5–fold more sequences per reported case (1.81% and 0.11% for HIC and LMICs, respectively) (**Table S3**). A moderate negative correlation between weekly sequencing percentages and reported COVID-19 incidence was observed (cases/100K pop., r^2^ = -0.52; *p*-value < 0.001), suggesting that countries with low incidence (**Fig. 1C; Fig. S3**) were able to sequence higher proportions of cases. Exceptionally, some countries, such as Denmark and the UK, faced high weekly COVID-19 incidence in late 2020 but were still able to maintain sequencing intensity >10% in most weeks (32% and 8% of total confirmed cases, respectively) (**Fig. 1A-B**; **Fig. S3**).

**Fig. 1.**
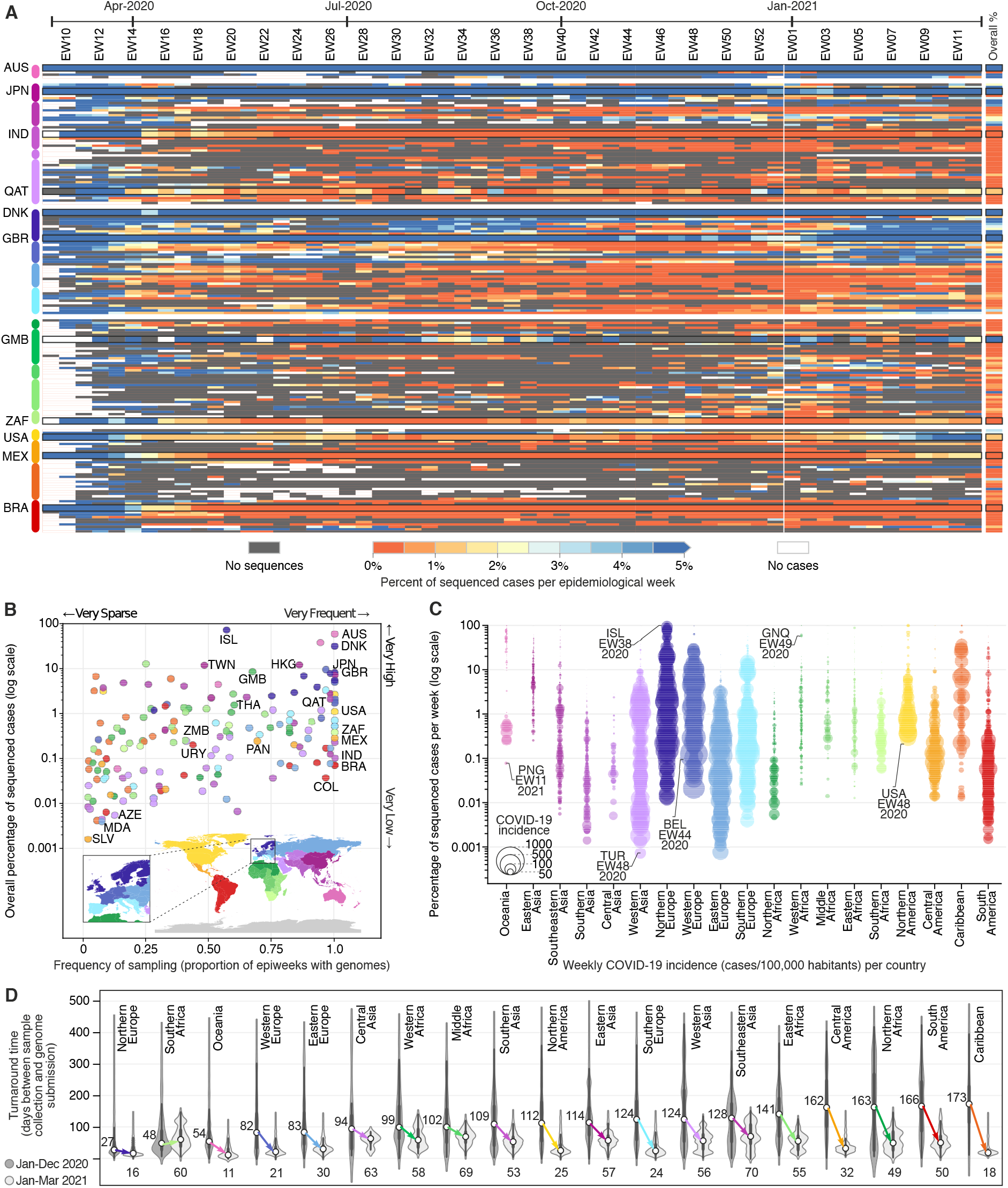
Disparities in SARS-CoV-2 global genomic surveillance. **(A)** Percentage of reported cases that were sequenced per country, per epidemiological week (EW), between February 23rd, 2020 and March 27th, 2021 (based on metadata submitted to GISAID up to May 30th, 2021). Updated numbers on sequence submissions and proportion of sequenced cases are available on the GISAID Submissions Dashboard at gisaid.org. **(B)** Frequency and overall percentage of sequenced cases per country. This plot summarizes the data shown in (A), where the x-axis shows the percentage of EWs with sequenced cases, and the y-axis displays the overall percentage of cases shown in the rightmost column of panel (A). **(C)** Percentage of cases sequenced per EW per country, per geographic region (classified according to the UNSD geoscheme). Each circle represents an EW with at least one sequenced case, and their diameters highlight the incidence (cases per 100,000 habitants), e.g. “ISL-EW38-2020” shows data from week 38 in 2020, in Iceland. **(D)** Distribution of turnaround times of genomes collected in different geographic regions, in 2020 and 2021. Countries are highlighted in panels of this figure using the ISO 3166-1 nomenclature.

Most countries in Africa and Asia, despite reporting low COVID-19 incidence, did not reach genomic surveillance levels similar to the Gambia (8.6%), Japan (7.3%), Hong Kong (12.3%), New Zealand (3.8%) and Australia (5.9%), which also experienced low COVID-19 incidences (**Fig. 1B-C**; **Fig. S3**). Likewise, sequencing of >0.5% of cases has not been achieved in most Latin American countries, particularly during periods of high incidence (**Fig. S3**). This finding is robust to under-ascertainment of reported cases due to more limited availability of diagnostic tests. Our study also revealed an absence of SARS-CoV-2 genomes in public databases from >20 LMICs; for some countries, the only available information on the diversity of circulating lineages has been obtained from travel-related infections sequenced abroad (**Fig. S2**). Overall, most countries did not achieve high or moderate percentages (0.1% to 1%) of sequenced cases each week of the pandemic (**Fig. 1**; **Fig. S3**).

We also described turnaround time (TAT; defined as the time in days between sample collection and genome submission) of SARS-CoV-2 genome sequencing across 19 geographic regions (**Fig. 1D**; see also (*11*)). On average, virus sequences were deposited in public databases 48 days after sample collection, but in 2021, following the detection of the Alpha VOC, efforts were made in nearly all geographic regions to decrease TAT, and provide faster responses (**Fig. 1D**; see **Fig. S4** for weekly changes in TAT across regions). Rapid generation and sharing of pathogen sequence data from regularly-collected samples is essential to maximize public health impact of genomic data (*12, 13*). The VOCs Alpha and Gamma, for example, reached up to 50% frequency within 2 to 3 months of their emergence in the U.K. and Manaus, respectively (*14, 15*). Thus, quick TAT is essential for the timely recognition and assessment of transmissibility potential of VOCs.

## Sampling strategies for rapid variant detection

We investigated the impact of genome sequencing intensity and TAT on the detection of SARS-CoV-2 lineages. Similar to what has been observed in the UK (*14*), the number of globally observed lineages correlates with the number SARS-CoV-2 genomes available per country (Pearson’s *r* = 0.96, p-value<0.0001) and the overall proportion of sequenced cases in each country (Pearson’s *r* = 0.48, p-value<0.0001) (**Fig. S5**). This implies that limited genome sequencing intensity may affect the identification and response to new viral lineages with altered epidemiological and antigenic characteristics. To investigate strategies for rapid variant detection, we simulated the impact of the percentage of sequenced cases and TAT on the reliable detection of previously-identified SARS-CoV-2 lineages using metadata from Denmark, which has one of the most comprehensive SARS-CoV-2 genome surveillance systems (see **Materials and Methods**). Because several countries have opportunistically selected samples for sequencing based on testing characteristics, e.g. spike gene target failures of a commonly-used PCR assay, or additional, often unspecified characteristics, such as imported cases or severe disease, we focused on analysing data collected prior to November 2020 (**Fig. S6**).

We assumed a recommended scenario of random sampling, whereby samples for virus genomic sequencing are selected independently of sample metadata such as age, sex, or clinical symptoms (*15*). When calculating the probability of detecting at least one genome of a rare lineage (0–5% prevalence) under different sequencing intensities, we found that sequencing at least 300 genomes per week is required to detect, with a 95% probability, a lineage that is circulating in a population at a weekly prevalence of 1%. For a weekly prevalence of 5%, this number decreases to 75 genomes per week (**Fig. 2A**). These figures are independent of outbreak and population size of a given location, and can only tell if a lineage is present, not how prevalent it is, and furthermore assumes representative sampling. On average, genome surveillance programmes in high income countries should be able to detect circulating virus lineages at 5% prevalence with maximum probability under the assumption of random sampling (**Fig. 2B; Table 1**). However, under a scenario of random sampling, low income countries that sequence an average of 9 genomes per week may miss a SARS-CoV-2 lineage circulating at up to 26% prevalence. This will present a substantial limitation to the lines of inquiry available to such countries from genome sequencing data (**Table 1)**. Within the range 0.05–5% sequences per case considered here, increasing sampling intensity and at a lesser extent reducing TAT strongly improves the rapid detection of viral lineages (**Fig. 2B**).

**Figure 2.**
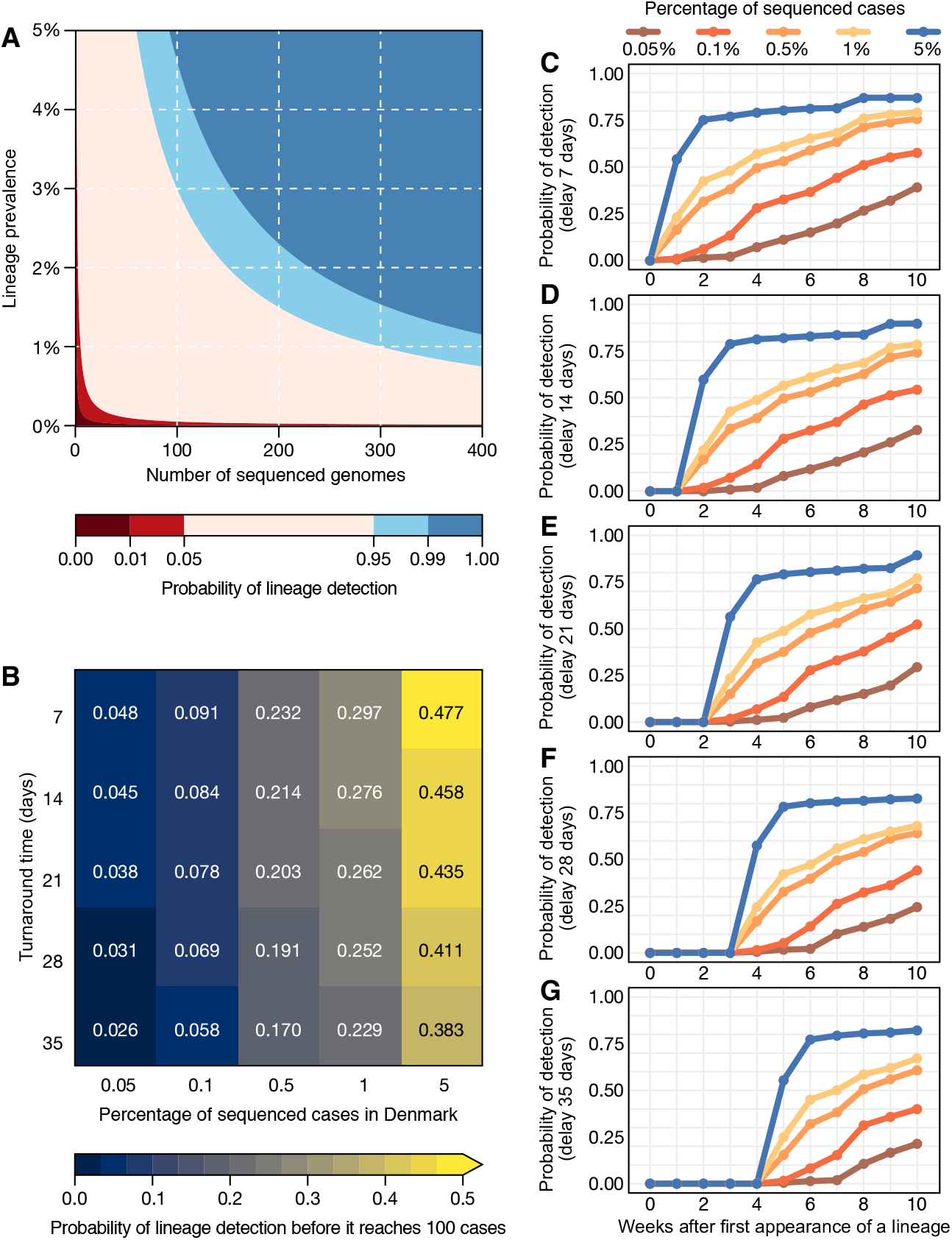
Detection of SARS-CoV-2 lineages under different genomic surveillance scenarios. (A) The probability of detecting at least one genome of a rare lineage under different sequencing regimes. (B) Relative importance of decreasing genome sequencing turnaround time (TAT) versus increasing sequencing percentage, measured as probability that a lineage found in simulated datasets was detected before it had reached 100 cases (described in **Fig. S6**). (C-G) Probability of lineage detection considering TATs of 7, 14, 21, 28 and 35 days.

**Table 1.**
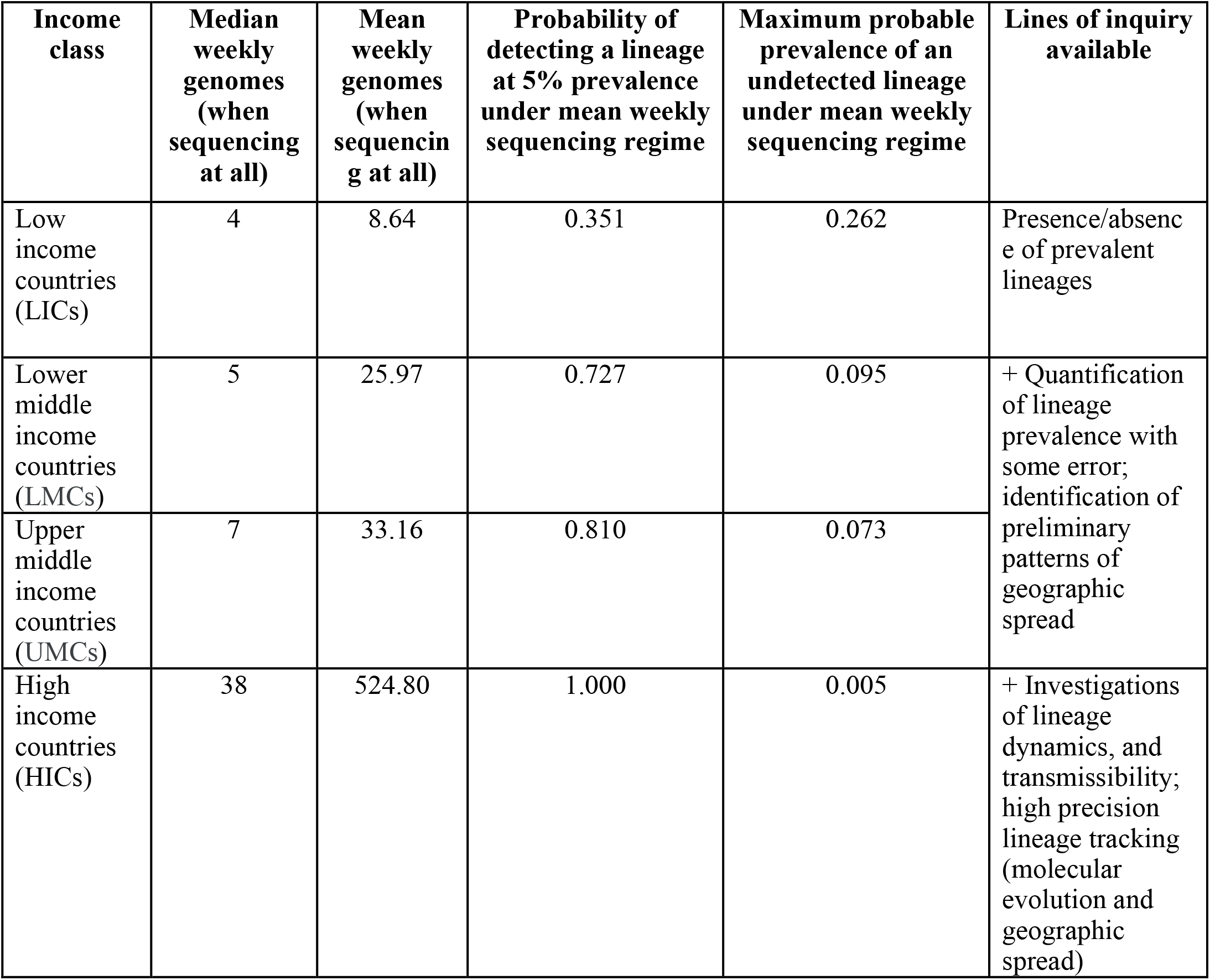
Empirical country sequencing capacities at different income levels and lines of inquiry enabled at each level. Countries at each income level have markedly different sequencing capacities, allowing for different degrees of epidemic resolution and lines of inquiry. Characteristics of each income class are shown in **Table S4**.

| **Income class** | **Median weekly genomes (when sequencing at all)** | **Mean weekly genomes (when sequencing at all)** | **Probability of detecting a lineage at 5% prevalence under mean weekly sequencing regime** | **Maximum probable prevalence of an undetected lineage under mean weekly sequencing regime** | **Lines of inquiry available** |
| --- | --- | --- | --- | --- | --- |
| Low income countries (LICs) | 4 | 8.64 | 0.351 | 0.262 | Presence/absence of prevalent lineages |
| Lower middle income countries (LMCs) | 5 | 25.97 | 0.727 | 0.095 | + Quantification of lineage prevalence with some error; identification of preliminary patterns of geographic spread |
| Upper middle income countries (UMCs) | 7 | 33.16 | 0.810 | 0.073 |  |
| High income countries (HICs) | 38 | 524.80 | 1.000 | 0.005 | + Investigations of lineage dynamics, and transmissibility; high precision lineage tracking (molecular evolution and geographic spread) |

Next, we simulated 25 scenarios with 100 replicates, in which we varied sampling frequency (from 0.05% to 5%) and TAT (from 35 to 7 days) to compute the probabilities of detecting at least one genome of a given lineage before the lineage reaches a cumulative size of 100 cases (**Fig. 2B**), using as “ground truth” a dataset from a well characterized setting (see **Materials and Methods** and **Fig. S6**). The simulated scenario shows that when sequencing percentages of 5% per week and turnaround times of 7 days are achieved in a given setting, there is a 48% probability of detecting a viral lineage before it reaches 100 cases randomly selected from the population. When the proportion of sequenced cases per week decreases by 100-fold, to 0.05%, the probability of the timely detection of a viral lineage before it reaches 100 cases decreases to 4.8% for turnaround times of 7 days, and further declines to 2.6% when turnaround time is 35 days (**Fig. 2B**).

For an optimistic scenario of 0.5% of sequenced cases (achieved by 69% of HICs and 23% of LMICs) and a turnaround time of 21 days (achieved by 14% of the HICs and 3% of the LMICs) (**Table S4**), we found a 20% probability of detecting a lineage before it reaches 100 cases. Throughout the pandemic, many countries reported weekly incidences as high as 100 cases per 100,000 inhabitants (**Fig. 1C, Fig. S3**). For example, in such a scenario of high incidence, for Manaus (2.2 million inhabitants, Amazonas state, Brazil), the 0.5% sequencing threshold would correspond to 11 randomly selected genomes per week. With a 21-day turnaround time, this would allow the detection of a given lineage with a 20% probability (**Fig. 2B**). For São Paulo (12.4 million inhabitants), this number increases to 62 genomes per week. For Brazil (212.6 million inhabitants), this would correspond to 1,063 weekly genomes selected from a random population of samples, in the above mentioned scenario of high incidence. Although the 0.5% ratio of sequenced cases per week in near real-time is a reasonable benchmark for SARS-CoV-2 genomic surveillance in over two thirds of high income country settings (**Table S4**), this often comes as a result of close coordination between diagnostic centers and well-funded, decentralized infrastructures to integrate sequencing data and sample-associated metadata (see e.g. (*16*)).

## Factors associated with genomic surveillance capacity

While many HICs were able to rely on previously established networks and laboratory infrastructure to perform molecular testing and sequencing (*17, 18*), many LMICs – including Brazil, South Africa, and India where three VOCs are believed to have emerged (*19*–*22*) - have faced additional challenges to rapid expansion of genomic surveillance (*18, 23, 24*). Pathogen genomics complements but often competes for limited resources with other aspects of pandemic response, for instance, surveillance and testing capacity, medical supplies, laboratory reagents, public health and social measures, vaccine development, and supplies (*25*). To investigate how socioeconomic factors can impact SARS-CoV-2 genomic surveillance response around the world, we explored the correlation between the percentage of sequenced COVID-19 cases in each country, and 20 country-level socioeconomic and health quality covariates (**Table S5**). We found that the percentage of sequenced cases are significantly associated with expenditure on research and development (R&D) per capita (r^2^=0.47, p-value<0.0001), GDP per capita (0.37, p-value<0.0001), socio-demographic index (0.31, p-value<0.001), and established influenza virus genomic surveillance capacity prior to the COVID-19 pandemic (0.30, p-value<0.001) (**Fig. 3; Table S6**).

**Figure 3.**
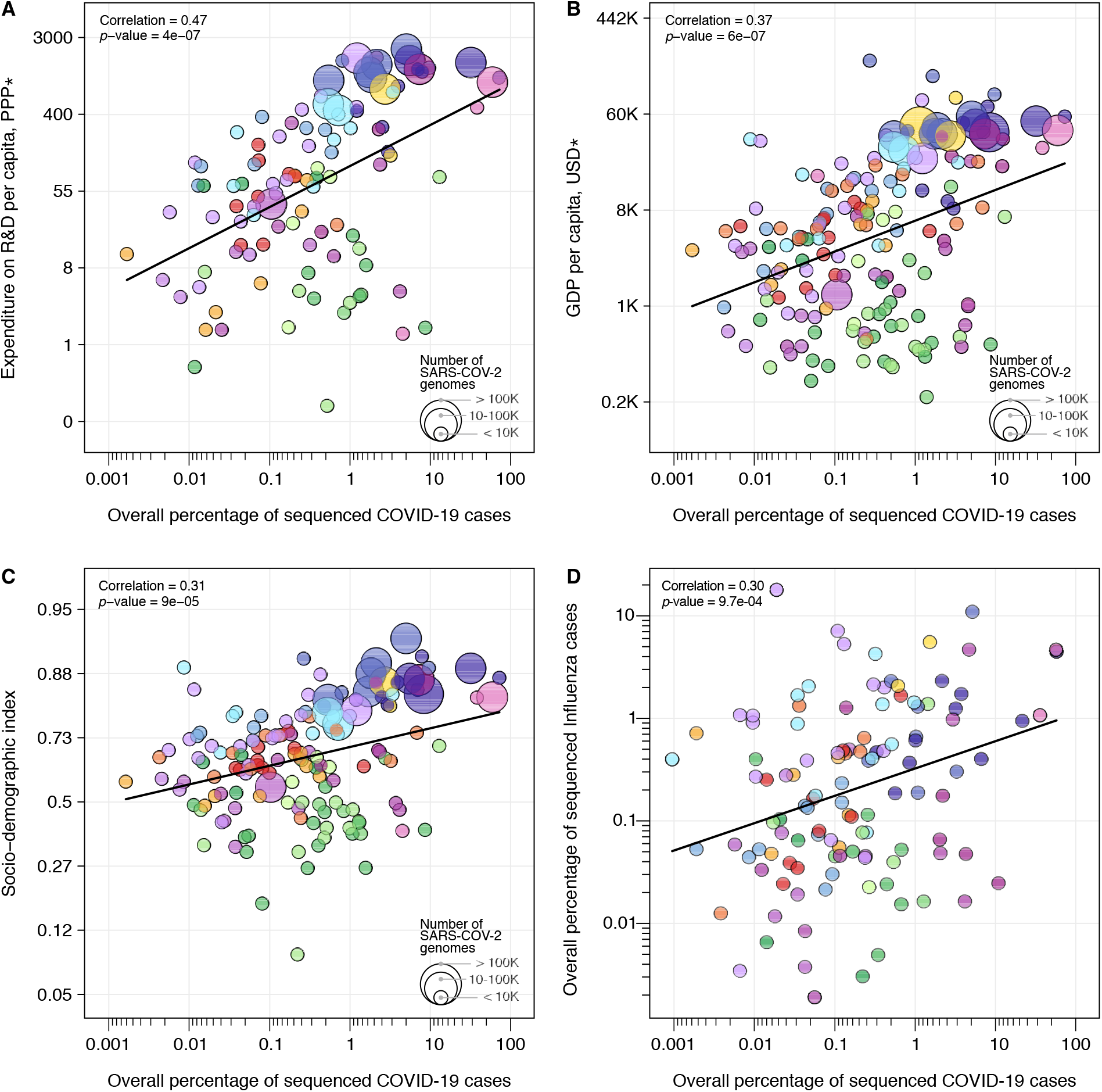
Case sequencing percentages and socioeconomic covariates. Covariates that show the highest correlation with the overall percentage of COVID-19 sequenced cases (during the period shown in Fig. 1A). (A) Expenditure on R&D per capita; (B) GDP per capita; (C) Socio-demographic index; (D) Overall percentage of influenza virus sequenced cases in 2019 (HA segment). For correlations between covariates and turnaround time, see **Fig. S7**. The colour scheme is the same as in Figure 1 and 2. Solid line shows the linear fit. *PPP = purchasing power parity, USD = US dollar 2005.

Before January 2020, only 67% (113 out of 167) of the countries that uploaded SARS-CoV-2 genomes to public databases had shared influenza virus genome sequences. When we compared breakdown by income class, we observed that the majority of UMCs (77%) and HICs (78%) sequencing SARS-CoV-2 had already reported influenza virus sequences in public databases up to 2019. For LICs and LMCs countries, this number drops to 39% and 54%, respectively, suggesting that many LICs and LMCs initiated genome sequencing programmes during the COVID-19 pandemic. While disparities in investment in national health, research, and development continue to impact the ability of countries to scale up genomic surveillance intensity (*6, 18, 26*), the uptake in genomic surveillance by many LMICs and the association of sequencing efforts with established genomic surveillance capacity provide an encouraging picture for future pandemic preparedness programmes.

When we explored correlations with mean turnaround time (**Table S7**), we found that universal health coverage (r^2^=-0.45, p-value<0.0001), healthcare access and quality index (−0.44, p-value<0.0001), socio-demographic index (−0.42, p-value<0.0001), and health expenditure per capita (−0.4, p-value<0.0001) are significantly correlated with mean turnaround times (**Fig. S7, Table S7**). Our results quantify only correlations between socioeconomic covariates, sequencing intensity, and turnaround time, and cannot be interpreted as causal. Future studies should focus on additional variables, such as training laboratory and bioinformatic personnel, costs associated with imported consumables, and shipment delays that may be exacerbated by border closures and travel restrictions (*6, 23, 24, 26, 27*). Other factors associated with delays in reporting VOCs include social and political stigma and perceived negative impact on travel when reporting potential VOCs, and concerns of having findings scooped and published by other researchers (*28*). Longer turnaround times are also expected in countries where virus genomics activities are focused on retrospective genomic studies to investigate SARS-CoV-2 reinfections (*29*), vaccine breakthrough infections (*30*), and past epidemic dynamics (*31, 32*).

## Conclusions

Strengthening pathogen genomic surveillance efforts worldwide, but particularly in LMICs, should be a global priority to improve pandemic preparedness. Our findings demonstrate that global SARS-CoV-2 genomic surveillance efforts are currently highly unbalanced, and contingent upon socioeconomic factors and pre-pandemic laboratory and surveillance capacity. Our results suggest that sequencing 0.5% of total confirmed cases, with a TAT below 21 days, could provide a benchmark for genomic surveillance studies targeting SARS-CoV-2 and future emerging viruses. Ongoing surveys to understand barriers to virus genome sequencing and sampling selection strategies will provide valuable information for future surveillance programmes. Implementation of metagenomic approaches for virus discovery followed by virus-genome specific sequencing approaches could help overcome existing limitations of molecular and syndromic surveillance strategies (*33*). Adoption of standardized protocols for representative genomic surveillance strategies (*15, 34*), rapid integration of sequence and sample-associated metadata, and collaboration between academia, public health laboratories and other stakeholders will be essential to maximize cost-effectiveness and public health impact of genomic surveillance. While a random sampling strategy may provide accurate information into SARS-CoV-2 variant emergence and frequency estimation, we note that genome sampling strategies should be considered pathogen- and question-specific (*15*). For example, non-random selection of samples stratified by disease severity may be required to identify genes or mutations associated with clinical outcomes.

Our findings call for strengthening equitable strategies that increase confidence in data sharing for improving global genomic surveillance (*28*). There are several global efforts underway to improve genomic sequencing capacities around the world, including the AFRO-Africa Centre for Disease Control, the Pan American Health Organization COVIGEN Network, South East Asian SARS-CoV-2 Genomics Consortium, and the ACT-A WHO Global Risk Monitoring Framework. These global efforts must be made to improve in-country genomic surveillance capacity and guarantee sustainable research funding for low and middle income countries. Improved pathogen surveillance at the human, animal and human-animal interfaces is also urgently needed (*35*). Retaining existing and expanding local capacity efforts acquired during the SARS-CoV-2 pandemic will be critical to contain and respond to the next “Disease X’’ (*35*).

## Data Availability

Data used in this study can be found in this GitHub repository: https://github.com/andersonbrito/paper_2021_metasurveillance

## Acknowledgments

We gratefully acknowledge the authors from the Originating laboratories responsible for obtaining the specimens, as well as the Submitting laboratories where the genomic data were generated and shared via GISAID, on which this research is based. An acknowledgement table can be found in Table S8 and at gisaid.org with set accession EPI_SET_20211008ez. We thank James Nokes, Isabella Lynette Ochola, and Sylvie Briand for their valuable comments. GD acknowledges Joshua Batson, whose work shared on Twitter (@thebasepoint) inspired the creation of Figure 2A. BHP and VS acknowledge the contribution of SARS-CoV-2 genomes by members of the Communicable Diseases Genomics Network of Australia.

## Author Contributions

Conception: AFB, NDG, NRF; Data acquisition: Danish Covid-19 Genome Consortium, COVID-19 Impact Project, Swiss SARS-CoV-2 Sequencing Consortium, Bulgarian SARS-CoV-2 sequencing group, Network for Genomic Surveillance in South Africa (NGS-SA), GISAID core curation team, GG, CNA, RTPL, MMS, PCR, CVFC, NSDS, SMS; Analysis: AFB, ES, GD, GWH, CCK, JH, SMS, SB, SF, MAS, GB; Interpretation: AFB, ES, GD, GWH, MUGK, JH, HT, GG, CNA, TdO, RTPL, SMS, SCH, OGP, CD, SB, SF, NDG, GB, NRF; Drafting: AFB, ES, GD, CCK, GB, NRF; Revising: AFB, ES, GD, GWH, MUGK, JH, HT, GG, CNA, LEM, CW, BPH, VS, NSZ, OM, HMB, TdO, RTPL, MMS, PCR, ATRV, FRS, RSA, IA, INI, IP, CVFC, NSDS, CG, SMS, DN, MP, MvK, SCH, ECS, OGP, CD, MAS, NDG, GB, NRF; Funding: NDG, NRF.

## Data and materials availability

All data and scripts used to generate figures and tables are available at https://github.com/andersonbrito/paper_2021_metasurveillance

## Supplementary Materials

Materials and Methods

Table S1–S8

Fig. S1–S7

