## Supplementary material for "Global disparities in SARS-CoV-2 genomic surveillance": Materials and Methods; Supplementary Materials

**This PDF file includes:**

Materials and Methods
Table S1–S8
Fig. S1–S7

### Materials and Methods

#### Genomic surveillance and epidemiological data

To obtain the percentage of sequenced cases for each country, per week and cumulative, we used metadata related to the "country of exposure" of genomes submitted to GISAID (*36*) up to May 30th, 2021, collected between epidemiological weeks (EWs) 9 of 2020 (February 23rd, 2020) and 12 of 2021 (March 27th, 2021). We obtained global daily COVID-19 case counts from Johns Hopkins University, Center for Systems Science and Engineering (CSSE) (http://github.com/CSSEGISandData/COVID-19), and population data from each country from the United Nations' Department of Economic and Social Affairs (*37*). Countries were grouped by income using the current classification by the World Bank (*38*). We calculated weekly percentages of COVID-19 cases sequenced per country by aggregating and dividing genome and case counts per EW, using a custom pipeline 'subsampler' (http://github.com/andersonbrito/subsampler).

#### Analysis of covariates correlated with genomic surveillance capacity

Covariates related to health systems were available from (*39*), GDP data were available from (*40*) and data on R&D expenditure per capita were available from (*41*). For the covariates from (*39*) we have selected their values for the year 2019, for GDP data from (*40*) for the year 2015, and for R&D expenditure we calculated country-wise means for the years 2013 through 2019. Influenza virus genomic data (HA segment) collected in 2019 were obtained from GISAID (*8*), and 2019 influenza death estimate data were downloaded from the IHME Global Burden of Disease Study 2019 (*39*). Correlations and covariate details are provided in **Table S5**. To calculate correlations, the percentage of sequenced cases was $\log_{10}$-transformed. Transformations applied to covariates are provided in **Table S6**, in column 'transformation'. For each covariate we have estimated a linear fit by applying a generalised linear model, regressing a (possibly, transformed, as indicated in **Table S6**) covariate onto the $\log_{10}$-transformed percentage of sequenced cases; *p*-values corresponding to the estimated slopes are available in Fig.s S3 and S4, column '*p*-value'.

#### Simulation of scenarios of genome sampling

As shown in Figure 1, Denmark has one of the most comprehensive genomic surveillance programs in this COVID-19 pandemic, sequencing around 35.6% of its reported cases up to May 16th, 2021 (260,183 cases and 92,592 genomes with >70% coverage; access date: May 30th, 2021) (*43*). In order to simulate the impact of the percentage of sequenced cases and the turnaround time (time between sample collection and genome submission) to reliably detect previously identified SARS-CoV-2 lineages in a country, we used metadata from genomes obtained by the Danish COVID-19 genome consortium, with collection dates between March and November 2020 (from EW 13 to EW 49) (*43*), to avoid potential distortions in lineage frequency caused by the preferential selection of variants for sequencing using S gene target failure (SGTF) data.

To evaluate the impact of delays on genome submission, based on the reported dates of sample collection, we generated lists of genomes with adjusted submission dates, to simulate turnaround times representing delays between 7 and 35 days (five weeks) between sample collection and genome submission. Considering the high percentage of sequenced cases per EW in Denmark (often above 20%), we produced several genome datasets simulating scenarios with different percentages of sequenced cases per EW (0.05%, 0.1%, 0.5%, 1% and 5%). By doing so we were able to simulate 25 scenarios (with 100 replicates each) with combinations of different turnaround times and percentage of sequenced cases, to assess how these two parameters may impact our ability (expressed as a probability) to detect circulating lineages. Specifically, we randomly sampled each column of the observed data (considered to be case counts across all circulating lineages) according to the targeted percentage of sequenced cases, which would become available after a given turnaround time. Each combination of percentage of sequenced cases and turnaround time yielded one table of genomes available across the EWs. This procedure was repeated 100 times to mitigate random sampling effects and to generate a probability of detection for each circulating lineage. Summarizing the 100 replicates led to detection probabilities for each lineage in each epi week.

**Fig. 2A** shows the probability of not drawing 0 from a Poisson distribution whose mean is the product of lineage prevalence and sequenced cases. In **Fig. 2B**, we show the computed probabilities of detection across simulation replicates, at a given sampling frequency and delay, which were able to have at least one detection of a given lineage before reaching a cumulative size of 100 cases in the full dataset without delays ("ground truth", see **Fig. S8**). **Figs. 2C-G** similarly map this out, but in time, asking how long it takes for a given lineage to be detected over time using the first instance of a lineage in the "ground truth" dataset as its emergence.

**Table S1.** Percentage of sequenced COVID-19 cases per country per epidemiological week (EW), between February 23rd, 2020 and March 27th, 2021 (based on metadata submitted to GISAID up to May 30th, 2021). The data shown here are the same used in Figure 1A to display weekly sequencing percentages. X = No cases; Code = ISO 3166-1 alpha-3; Income category = income category, according to the World Bank classification; Frequency of sampling = Proportion of weeks with at least one genome.

*[Available as a separate Excel file]*

**Table S2.** List of countries that mostly relied on other countries to get their COVID-19 cases sequenced.

*[Available as a separate Excel file]*

**Table S3.** Total number of sequenced SARS-CoV-2 genomes between February 23rd, 2020 and March 27th, 2021 (based on metadata submitted to GISAID up to May 30th, 2021), number of COVID-19 cases, and overall percentage of sequenced cases, per income category, according to the World Bank classification (year: 2019).

| Income category | Total genomes | Total cases | Overall percentage of sequenced cases |
|---|---|---|---|
| High income | 1,182,367 | 65,387,757 | 1.81% |
| Low-mid income | 70,164 | 61,202,215 | 0.11% |

**Table S4.** Key surveillance characteristics with a split by income class. We provide summary statistics of observed surveillance characteristics for each group of countries, defined by their income class. HIC - high income class, UMC - upper middle income class, LMC - low middle income class, LIC - lower income class, non-HIC - combined UMC, LMC and LIC.

| | **Surveillance intensity** | | **Timeliness** | | **Regularity** | |
|---|---|---|---|---|---|---|
| Income class | Overall percentage of sequenced cases >= 0.5% | Overall percentage of sequenced cases < 0.5% | Genomes submitted with turnaround time <=21 days | Genomes submitted with turnaround time > 21 days | Countries sequencing genomes in >=75% of the weeks | Countries sequencing genomes in <75% of the weeks |
| LIC | 0.41 | 0.59 | 0.00 | 1.00 | 0.00 | 1.00 |
| LMC | 0.22 | 0.78 | 0.05 | 0.95 | 0.20 | 0.80 |
| UMC | 0.18 | 0.82 | 0.02 | 0.98 | 0.29 | 0.71 |
| HIC | 0.69 | 0.31 | 0.14 | 0.86 | 0.59 | 0.41 |
| non-HIC* | 0.23 | 0.77 | 0.03 | 0.97 | 0.21 | 0.79 |

**Table S5.** Typical country profiles characterised by covariates. We provide typical values of covariates which characterise capacity and coordination abilities for each group of countries, linked to their income level. HIC - high income country, UMC - upper middle income country, LMC - low middle income country, LIC - low income country.

| Covariate | HIC | UMC | LMC | LIC | Covariate name | Covariate description |
|---|---|---|---|---|---|---|
| gdp | 35556 | 5280 | 1478 | 416 | GDP per capita | GDP per capita |
| erd | 732 | 86 | 19 | 5 | Expenditure on R&D per capita | Expenditure on R&D per capita in PPP (purchasing power parity dollars) |
| he_cap | 2941 | 1019 | 311 | 90 | Health expenditure (per capita) | The variable is health expenditure per capita taken from FGH April 2019, in 2018 USD |
| sdi | 0.83 | 0.68 | 0.53 | 0.33 | Socio-demographic Index | A measure of development estimated via principal component analysis using log-transformed LDI, TFR (ages 25+), and education years per capita over age 15 |
| fluprop | 1.84 | 0.53 | 0.16 | 0.11 | Proportion of sequenced Flu cases in 2019 | Genomic surveillance capacity |
| edu_gini_mat | 0.13 | 0.17 | 0.30 | 0.52 | Education Relative Inequality (Gini), maternal | Education Relative Inequality (Gini), maternal |
| gallup_neg_exp_index | 27 | 29 | 28 | 31 | Gallup: Negative Experience Index | Negative Experience Index estimated via the Gallup World Poll surveys |
| universal_health_coverage | 87 | 71 | 56 | 42 | Universal health coverage | Coverage of universal health coverage tracer interventions for prevention and treatment services, percent; created for GBD 2015 SDGs paper. |
| health_worker_density | 296 | 129 | 56 | 22 | Health worker density | Number of employed health workers (of any specialty) per 10,000 population |
| hospital_beds_per1000 | 4.19 | 2.96 | 1.81 | 0.66 | Hospital Beds (per 1000) | Hospital beds per 1000 people |
| ifd_coverage_prop | 0.99 | 0.97 | 0.82 | 0.66 | In-Facility Delivery (proportion) | Percent of women giving birth in a health facility |
| occ_professional | 0.15 | 0.10 | 0.06 | 0.04 | Occupation Professionals | The proportion of the employed population ages 15- |

| | | | | | | |
|---|---|---|---|---|---|---|
| | | | | | | 69 working as professionals (according to ISCO classifications) |
| pharmacists_pc | 14 | 6 | 3 | 1 | Pharmacists per capita | Number of employed pharmacists and pharmaceutical assistants per 10,000 population |
| physicians_pc | 29 | 17 | 8 | 2 | Physicians per capita | Number of employed medical doctors per 10,000 population |
| prop_urban | 0.42 | 0.35 | 0.33 | 0.25 | Urbanicity | Urbanicity |
| haqi | 86 | 65 | 46 | 30 | Healthcare access and quality index | Healthcare access and quality index |

135 **Table S6.** Correlations of country-level covariates with the percentage of sequenced COVID-19 cases. 'Transformation' column denotes the transformation applied to the corresponding covariate before assessing the correlation; the *p*-value column shows significance of the slope in a linear model.

| Covariate | Correlation | Transformation | *p*-value | Covariate name | Covariate description |
|---|---|---|---|---|---|
| erd | 0.47 | log | 4E-07 | Expenditure on R&D per capita | Expenditure on R&D per capita in PPP (purchasing power parity dollars) |
| av_gdp | 0.37 | log | 6E-07 | GDP per capita | GDP per capita |
| frac_oop_hexp | -0.35 | no | 9E-06 | Fraction of OOP Health Expenditure | Fraction of out-of-pocket health expenditure out of total health expenditure, from FGH April 2019 |
| sdi | 0.31 | logit | 9E-05 | Socio-demographic Index | A measure of development estimated via principal component analysis using log-transformed LDI, TFR (ages 25+), and education years per capita over age 15 |
| fluprop | 0.30 | log | 9E-04 | Percentage of sequenced Flu cases in 2019 | Genomic surveillance capacity |
| anc1_coverage_prop | 0.28 | logit | 6E-04 | Antenatal Care (1 visit) Coverage (proportion) | Proportion of pregnant women receiving any antenatal care from a skilled provider |
| he_cap | 0.28 | log | 6E-04 | Health expenditure (per capita) | The variable is health expenditure per capita taken from FGH April 2019, in 2018 USD |
| health_worker_density | 0.28 | log | 6E-04 | Health worker density | Number of employed health workers (of any specialty) per 10,000 population |
| occ_professional | 0.27 | no | 8E-04 | Occupation Professionals | The proportion of the employed population ages 15-69 working as professionals (according to ISCO classifications) |

| | | | | | |
|---|---|---|---|---|---|
| universal_health _coverage | 0.25 | no | 3E-03 | Universal health coverage | Coverage of universal health coverage tracer interventions for prevention and treatment services, percent; created for GBD 2015 SDGs paper. |
| haqi | 0.24 | no | 3E-03 | Healthcare access and quality index | Healthcare access and quality index |
| hospital_beds_pe r1000 | 0.22 | log | 8E-03 | Hospital Beds (per 1000) | Hospital beds per 1000 people |
| pharmacists_pc | 0.21 | log | 8E-03 | Pharmacists per capita | Number of employed pharmacists and pharmaceutical assistants per 10,000 population |
| edu_gini_mat | -0.2 | logit | 2E-02 | Education Relative Inequality (Gini), maternal | Education Relative Inequality (Gini), maternal |
| gallup_neg_exp_ index | -0.19 | no | 2E-02 | Gallup: Negative Experience Index | Negative Experience Index estimated via the Gallup World Poll surveys |
| contra_demand_ satisfied | 0.18 | no | 3E-02 | Demand for contraception satisfied with modern methods | Proportion of women with a demand for contraception that are using a modern method |
| ifd_coverage_pr op | 0.17 | logit | 4E-02 | In-Facility Delivery (proportion) | Percent of women giving birth in a health facility |
| physicians_pc | 0.12 | log | 1E-01 | Physicians per capita | Number of employed medical doctors per 10,000 population |
| war_rate | -0.11 | logit | 2E-01 | Mortality Rate Due to War Shocks (per 1 person) | Mortality rate per one person due to war and terrorism (cause_id: 945); updated for GBD 2016 definition of war and terrorism |
| prop_urban | 0.03 | no | 7E-01 | Urbanicity | Urbanicity |

**Table S7.** Correlations of country-level covariates with the mean turnaround time. 'Transformation' column denotes the transformation applied to the corresponding covariate before assessing the correlation; the *p*-value column shows significance of the slope in a linear model.

| Covariate | Correlation | Transformation | *p*-value | Covariate name |
|---|---|---|---|---|
| universal_health_coverage | -0.45 | no | 2E-08 | Universal health coverage |
| haqi | -0.44 | no | 4E-08 | Healthcare access and quality index |
| sdi | -0.42 | logit | 3E-07 | Socio-demographic Index |
| he_cap | -0.4 | log | 1E-06 | Health expenditure (per capita) |
| health_worker_density | -0.37 | log | 4E-06 | Health worker density |
| av_gdp | -0.34 | log | 9E-06 | GDP per capita |
| edu_gini_mat | 0.33 | logit | 6E-05 | Education Relative Inequality (Gini), maternal |
| hospital_beds_per1000 | -0.33 | log | 5E-05 | Hospital Beds (per 1000) |
| erd | -0.32 | log | 1E-03 | Expenditure on R&D per capita |
| occ_professional | -0.31 | no | 2E-04 | Occupation Professionals |
| ifd_coverage_prop | -0.3 | logit | 3E-04 | In-Facility Delivery (proportion) |
| physicians_pc | -0.3 | log | 3E-04 | Physicians per capita |
| pharmacists_pc | -0.29 | log | 5E-04 | Pharmacists per capita |
| anc1_coverage_prop | -0.24 | logit | 5E-03 | Antenatal Care (1 visit) Coverage (proportion) |
| contra_demand_satisfied | -0.23 | no | 7E-03 | Demand for contraception satisfied with modern methods |
| prop_urban | -0.2 | no | 2E-02 | Urbanicity |

| | | | | |
|---|---|---|---|---|
| fluprop | -0.18 | log | 5e-02 | Percentage of sequenced Flu cases in 2019 |
| gallup_neg_exp_index | 0.16 | no | 6e-02 | Negative Experience Index estimated via the Gallup World Poll surveys |
| war_rate | 0.16 | logit | 6E-02 | Mortality Rate Due to War Shocks (per 1 person) |
| frac_oop_hexp | 0.15 | no | 7E-02 | Fraction of OOP Health Expenditure |

**Table S8.** GISAID acknowledgment Table (also available at gisaid.org with set accession EPI_SET_20211008ez).

*[Available as a separate TSV file]*

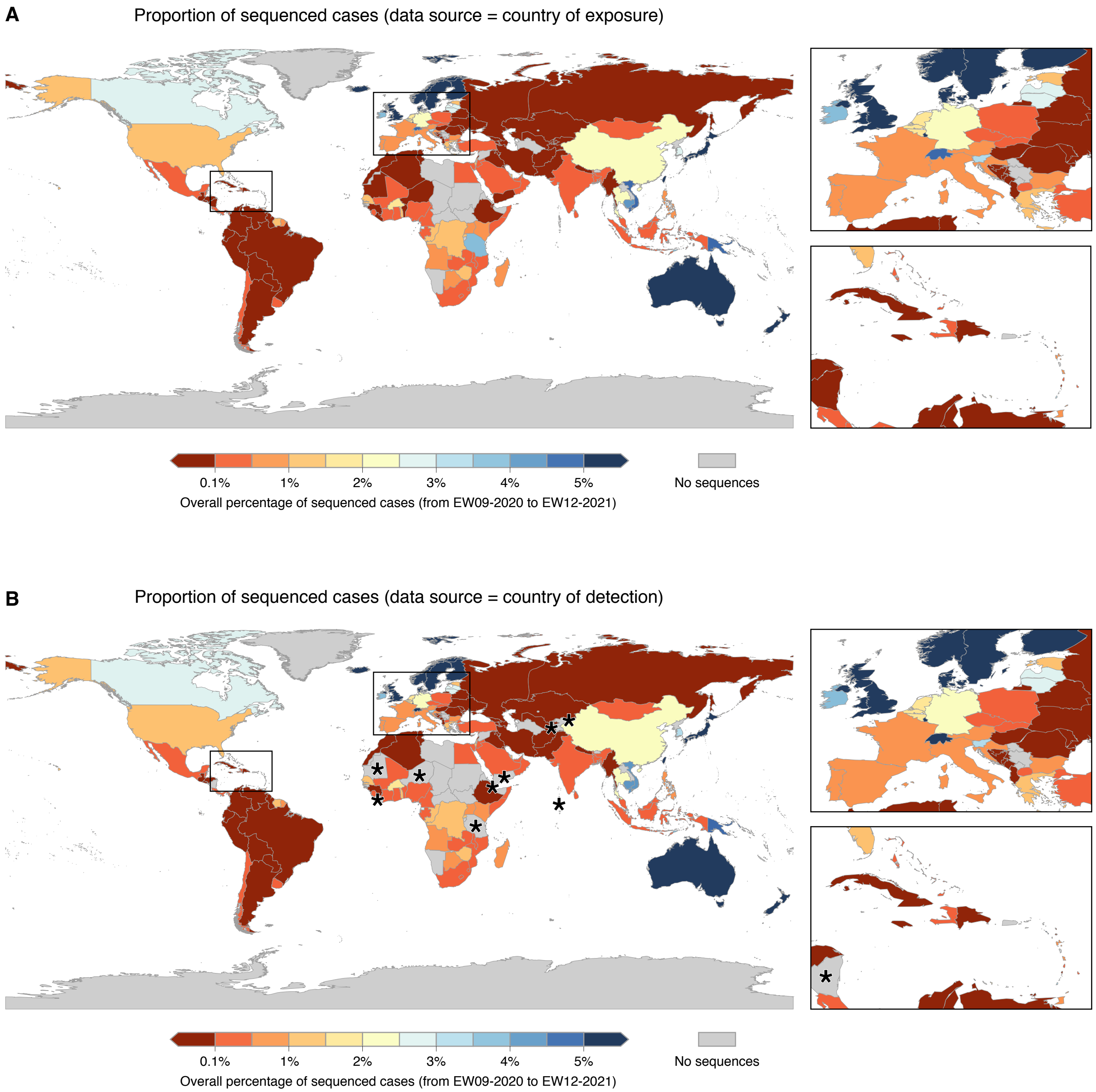


**Fig. S1.** Overall percentage of sequenced cases per country, between EW09 of 2020 and EW12 of 2021. The data shown here are the same used in Figure 1 to display weekly sequencing percentages. (A) Sequencing percentages observed when “country of exposure” is used as data source for defining the geographic origin of genomes, to reflect the locations where infections started (instead of where cases were detected). (B) Sequencing percentages observed when “country of sampling” is used as data source for defining the geographic origin of genomes, to reflect the locations where the infections were detected and where the cases were sequenced. As shown, genomic surveillance in some countries (marked with *, asterisks) rely entirely on data obtained abroad, generated from travel cases.

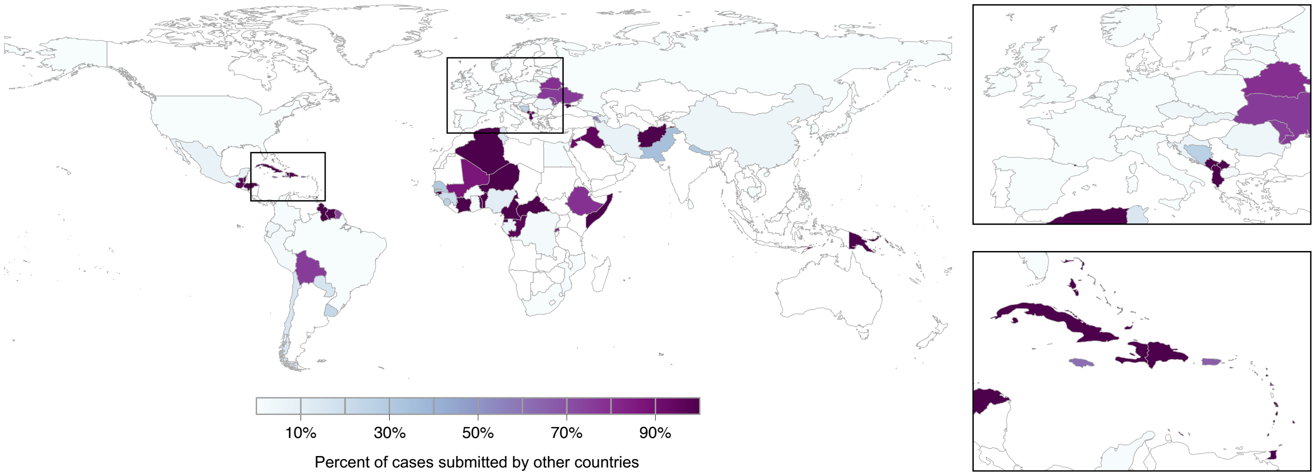


**Fig. S2.** Countries that rely mostly on other countries' capacity for genome sequencing and submission. Countries that rely on external resources are highlighted with shades of purple, based on the percentage of their cases that were sequenced and submitted by other countries.

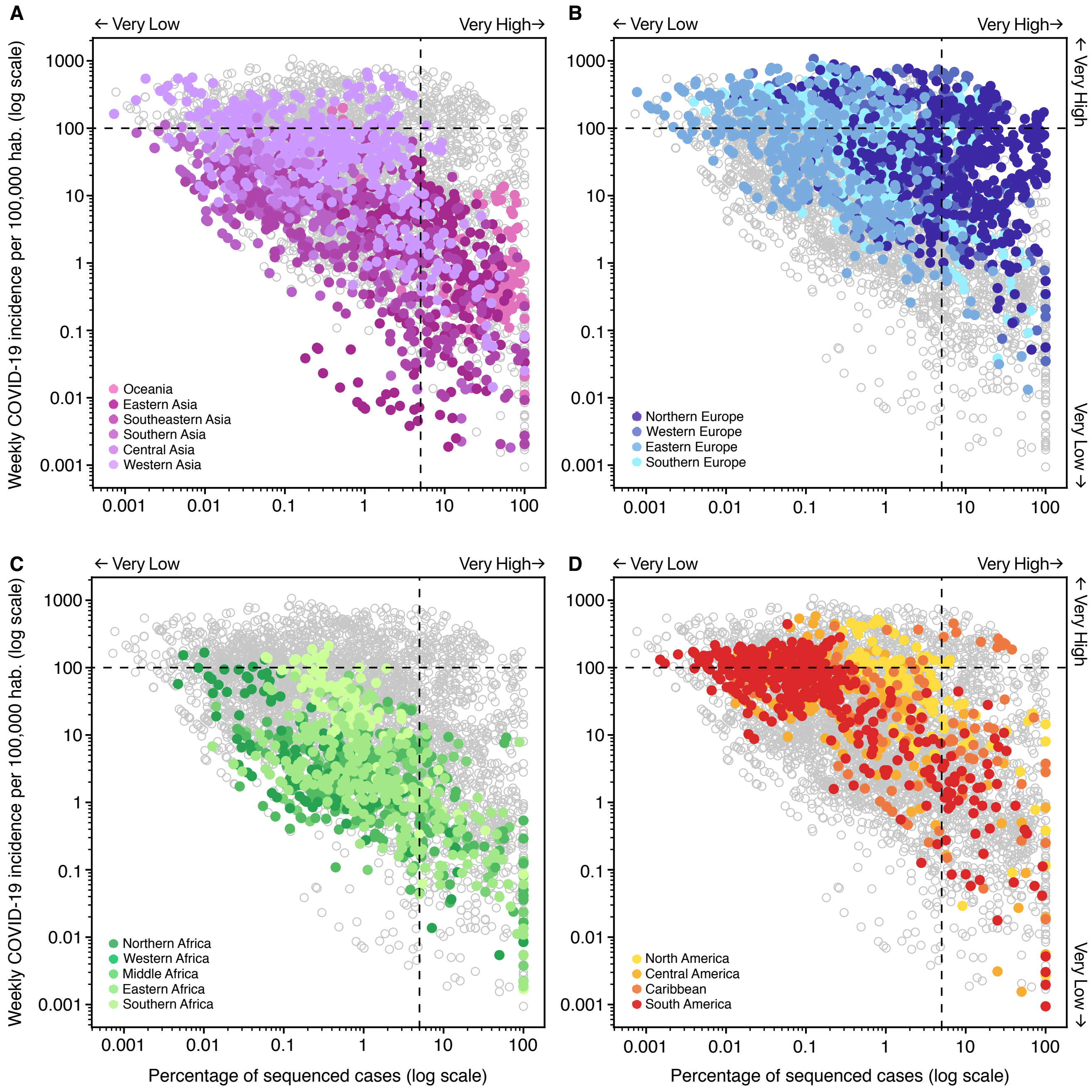


**Fig. S3.** Correlation between weekly COVID-19 incidence per 100,000 habitants, and percentage of sequenced cases in **(A)** Oceania & Asia, **(B)** Europe, **(C)** Africa and **(D)** the Americas, using the same data displayed in Figure 1, where each point represents an epidemiological week in a country. Vertical dashed lines represent the threshold of 5% sequenced cases, while the horizontal line marks 100 cases per 100,000 habitants (high COVID-19 incidence).

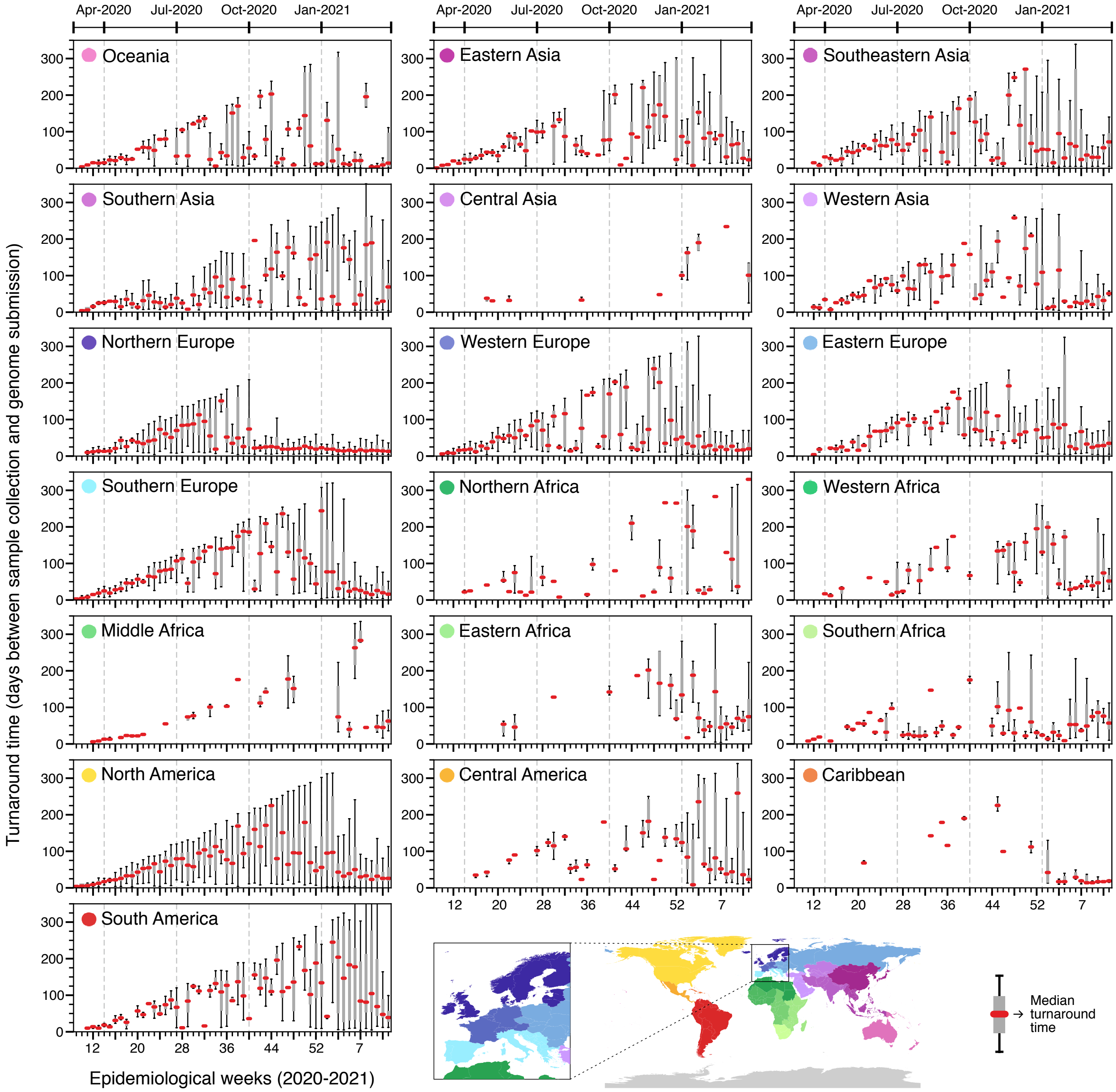


**Fig. S4. Turnaround time across geographic regions**. Delays between sample collection and genome submission across epidemiological weeks (turnaround time) in different regions, between February 23rd, 2020 and March 27th, 2021, based on metadata submitted to GISAID up to May 30th, 2021.

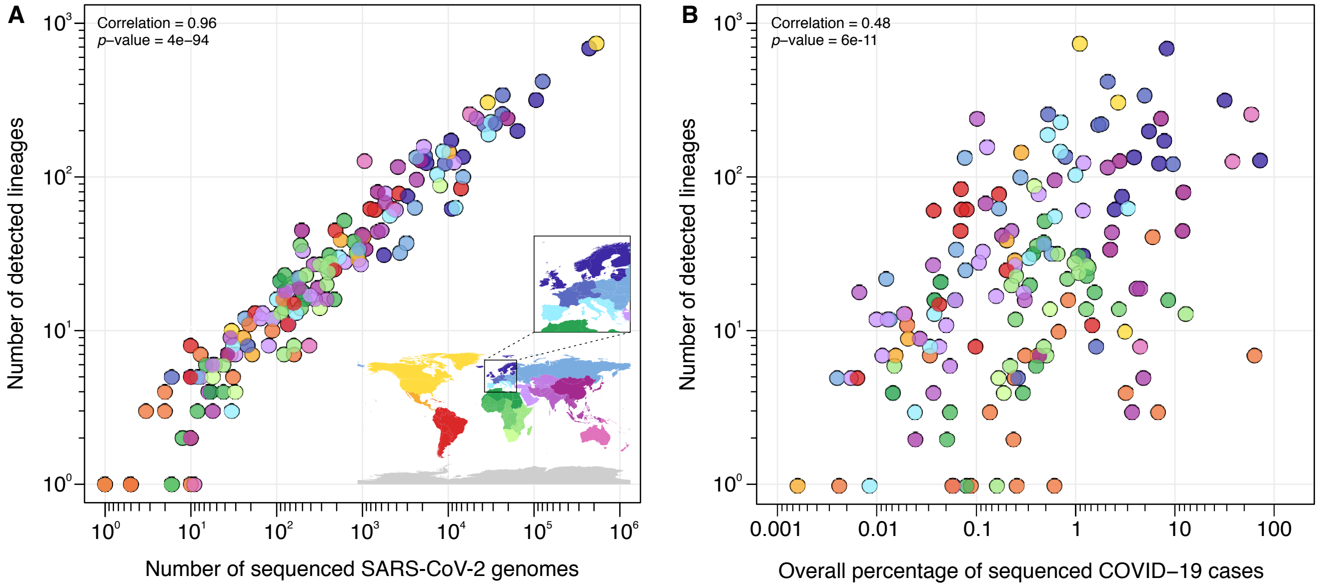


**Fig. S5.** Correlation between $\log_{10}$-transformed number of detected lineages and $\log_{10}$-transformed **(A)** number of sequenced genomes and **(B)** percentages of sequenced cases per country.

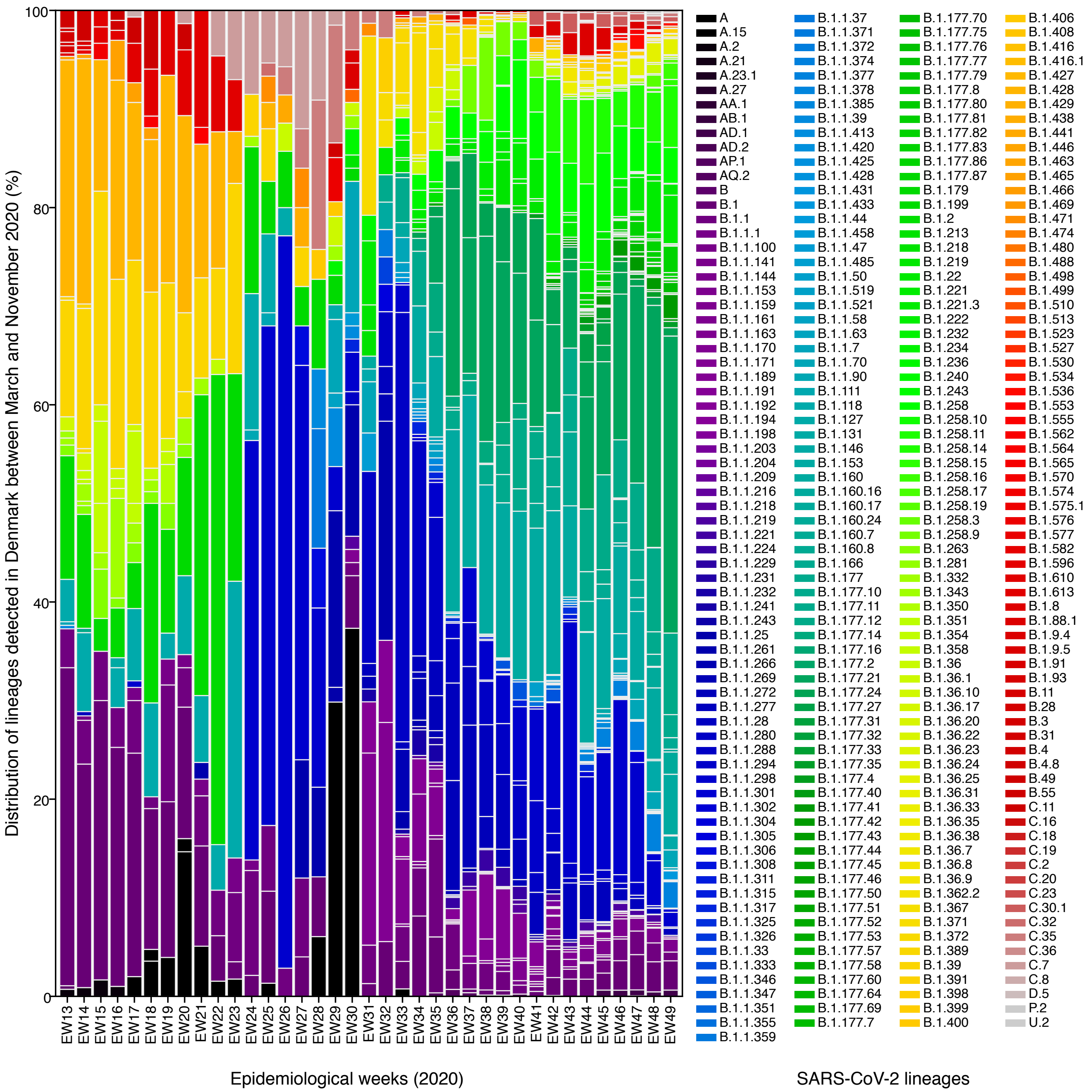


**Fig. S6.** Relative frequency of lineages detected in Denmark between epi weeks 13 and 49 (grouped by collection dates). In this period the country sequenced more than 20% of its reported cases, on average, and this dataset was used as the ‘ground truth’ for the simulations of probabilities of lineage detection shown in Figure 2B-G.

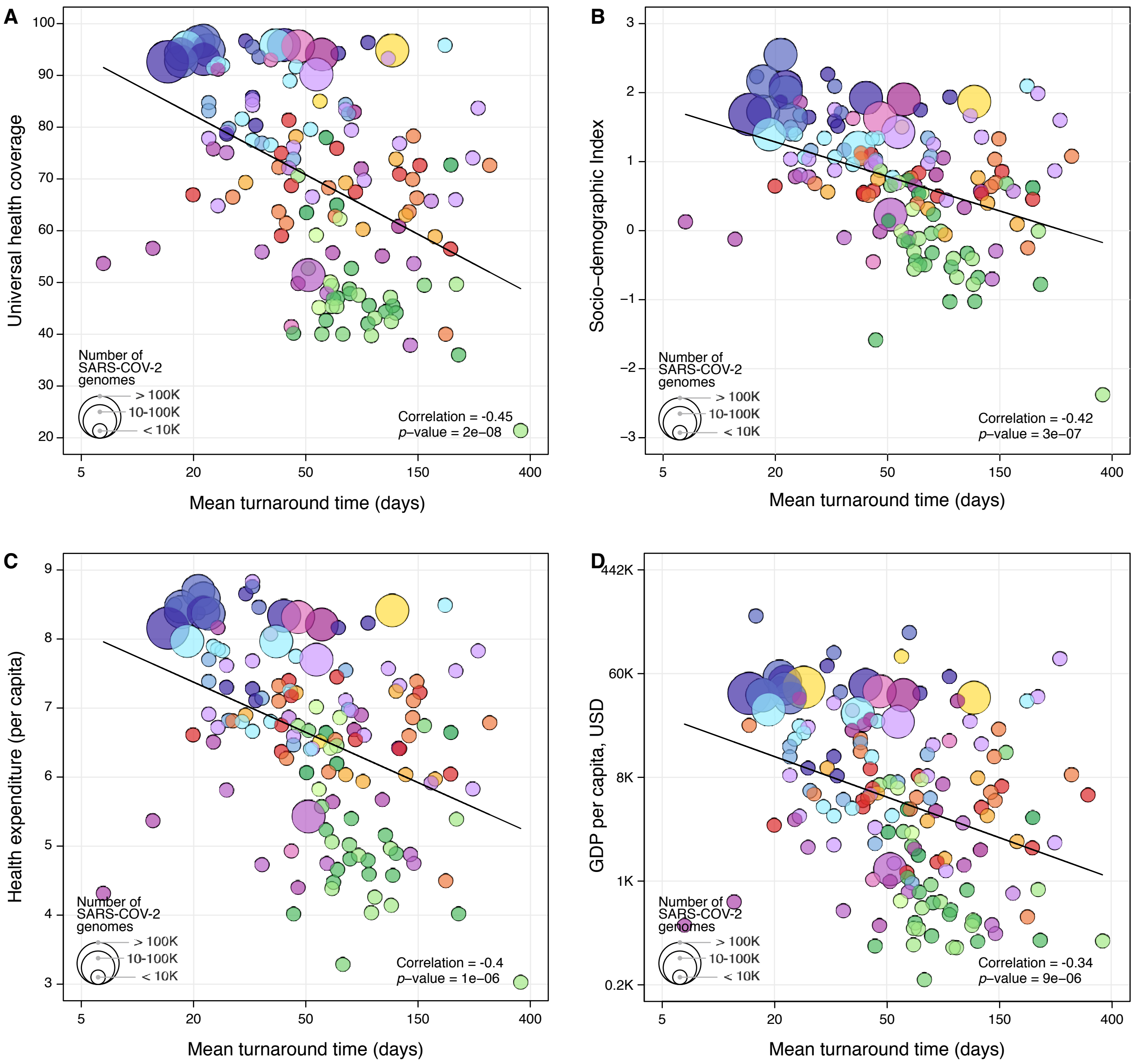


**Fig. S7.** Covariates that show the highest negative correlation with the mean turnaround time. **(A)** Universal health coverage; **(B)** Socio-demographic Index; **(C)** Health expenditure (per capita); **(D)** GDP per capita, in USD. The colour scheme of geographic regions is the same used in Figure 1. A solid line shows the linear fit in each figure.
